# SARS-CoV-2 alpha variant: is it really more deadly? A population level observational study

**DOI:** 10.1101/2021.08.17.21262167

**Authors:** Candace Makeda Moore, Ruslan Sergienko, Ronen Arbel

## Abstract

**Background:** In 2021 a new variant of SARS-CoV-2, which came to be called the alpha variant, spread around the world. There were conflicting reports on this COVID-19 variant strain’s potentially increased lethality. In Israel, this strain became predominant in a very short time period.

**Methods:** COVID-19 mortality and case fatality rates were examined in Israel in terms of weekly and cumulative numbers.

**Results:** COVID-19 case fatality rates in Israel rose quickly at the beginning of the pandemic and peaked in May 2020. The highest crude mortality came later in the second and third waves, but case the case fatality rates did not rise in 2021 with the increasing dominance of the alpha variant.

**Conclusions:** Based on the results of examining case-fatality and mortality rates, we concluded that while the alpha variant of the virus raised mortality, in line with the fact that it is more infectious than wild-type, once this strain was caught by patients in Israel, it was not more likely to kill them than the original strain

## Introduction

In 2021 new variants of SARS-CoV-2 spread around the globe including alpha, beta, gamma, delta, eta and lambda. New variants are of special concern if they are more infectious or more deadly once caught, but a clear agreement about how to determine which of these possibilities has driven up cases in some regions has not yet formed., spread across Europe (1) and beyond. The variant is now referred to as the alpha variant, first termed B.1.1.7, was first identified in the UK, it rapidly became a global phenomenon (1)(2). By mid-January, this variant had been reported in twelve states of the United States of America (3). During the same month of January, the alpha variant became the dominant strain in Israel, making up over 90% of cases (4). Israel intensified surveillance and prioritized rapid vaccination (4). The public health implications of the new strain were dependent on how it would affect the population; however, there was some controversy. The alpha variant strain was reported as more transmissible (5) and perhaps more deadly (2, 6-8). However, the evidence that alpha variant was deadlier than the wild type is controversial since a study of hospitalized patients showed that the variant did not produce more severe disease or a higher death rate (9) than the wild type. A better understanding of the true implications of this strain of virus is critical for healthcare workers around the globe, as uncertainty has been a source of anxiety for frontline care providers, especially in countries like Saudi Arabia where many healthcare workers are expatriates for whom this specific strain has been a source of anxiety (10). Examination of real-world epidemiological outcomes can also provide an exemplary framework to evaluate new variants as they emerge. In Israel, COVID-19 testing and care are freely available to all residents. Therefore, the strain switch provided a natural experiment for the effects of different strains. Because of the differing evidence on whether the alpha variant strain was deadlier than the wild-type, our objective was to analyze the Israeli data on COVID-19 to determine whether the alpha variant produced a higher fatality rate.

## Methods

The primary outcomes were the fatality rates of COVID-19 over time as the predominant strain changed from the wild type to the alpha variant strain. We analyzed the data published by the Israel Ministry of Health from March 1st, 2020, until February 28th, 2021, when massive vaccination started to affect mortality significantly. The crude mortality rate was calculated by summing deaths divided by Israel’s total population. Case-fatality rates were calculated weekly to minimize weekend under-reporting biases. We calculated the cumulative case-fatality rate from July 2020 to minimize the bias from the learning curve of the Israeli health system to cope with COVID-19 when it began.

To ensure statistical validity of our results, we used time periods before and after the emergence of the Alpha variant in Israel and compared the case fatality ratios with the Fisher’s exact test. We included the results from December 7th to December 21st compared to December 27^th^ to January 10^th.^

## Results

Case fatality rates rose quickly at the beginning of the pandemic and peaked in May 2020. The highest crude mortality came later in the second and third waves, but case fatality was lower, as illustrated in Figure 1.

**Figure One:**
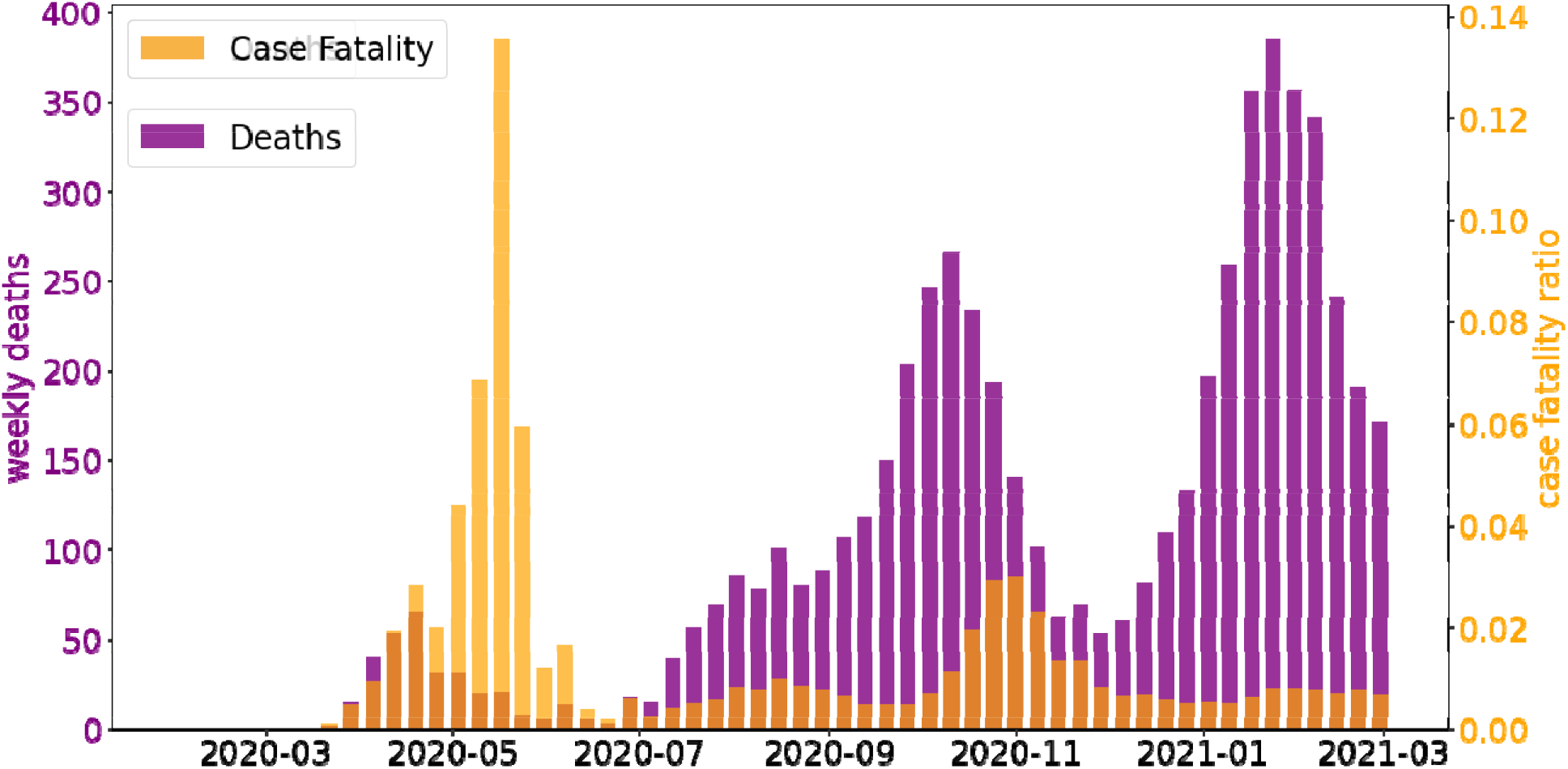
Weekly case fatality and overall mortality rates in Israel

The first cases of alpha variant were discovered in Israel on December 22nd, 2020, and the strain became dominant in January 2021. An examination of the cumulative case-fatality rates shows stabilization. The case fatality rates did not rise in 2021 with the increasing dominance of the alpha variant. Cumulative case-fatality rates from July 2020 until the end of March 2021 actually decreased from 1.25 to 0.74 percent. An examination of cumulative mortality rates shows stabilization, as illustrated in figure two.

**Figure Two:**
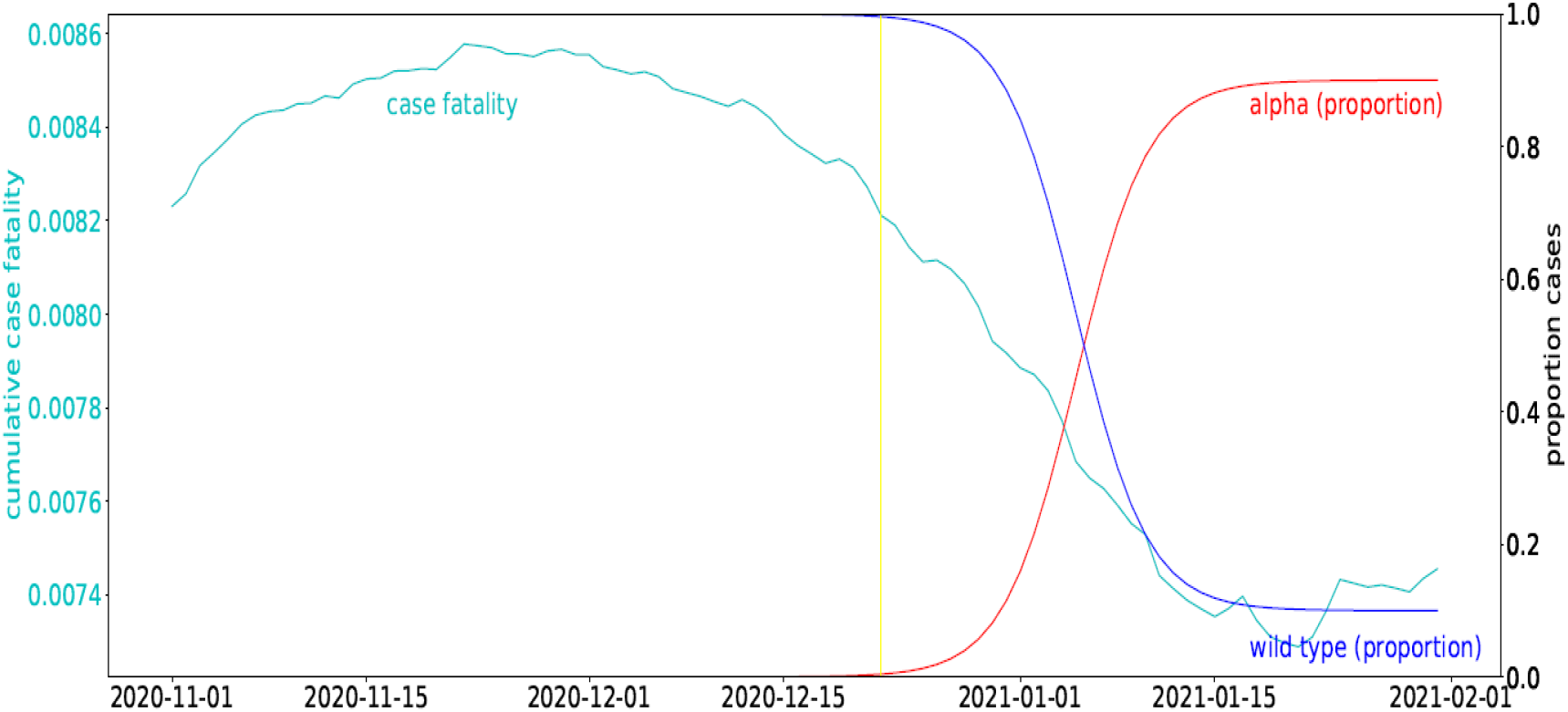
Cumulative case fatality rates from November 2020 until the end of January 2021. The horizontal yellow line marks the date the alpha variant first appeared in Israel. Variant proportions are illustrated.

The Fisher exact test of case fatality rates for periods before and after the emergence of alpha produced the Fisher exact test statistic value of 0.0268. The result is significant at p < .05. Data for cases and deaths in November, December and January are available in table one.

**Table 1:**
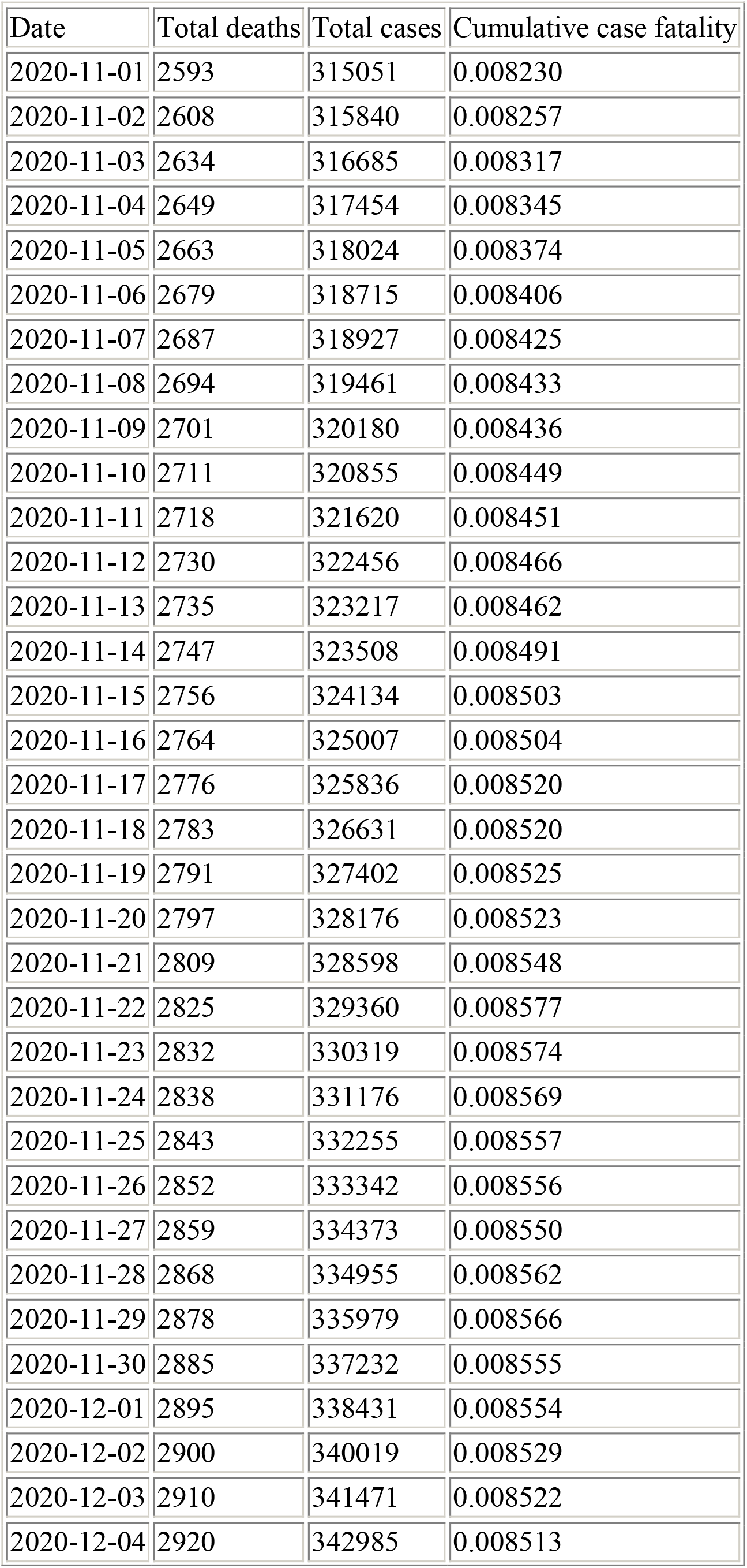
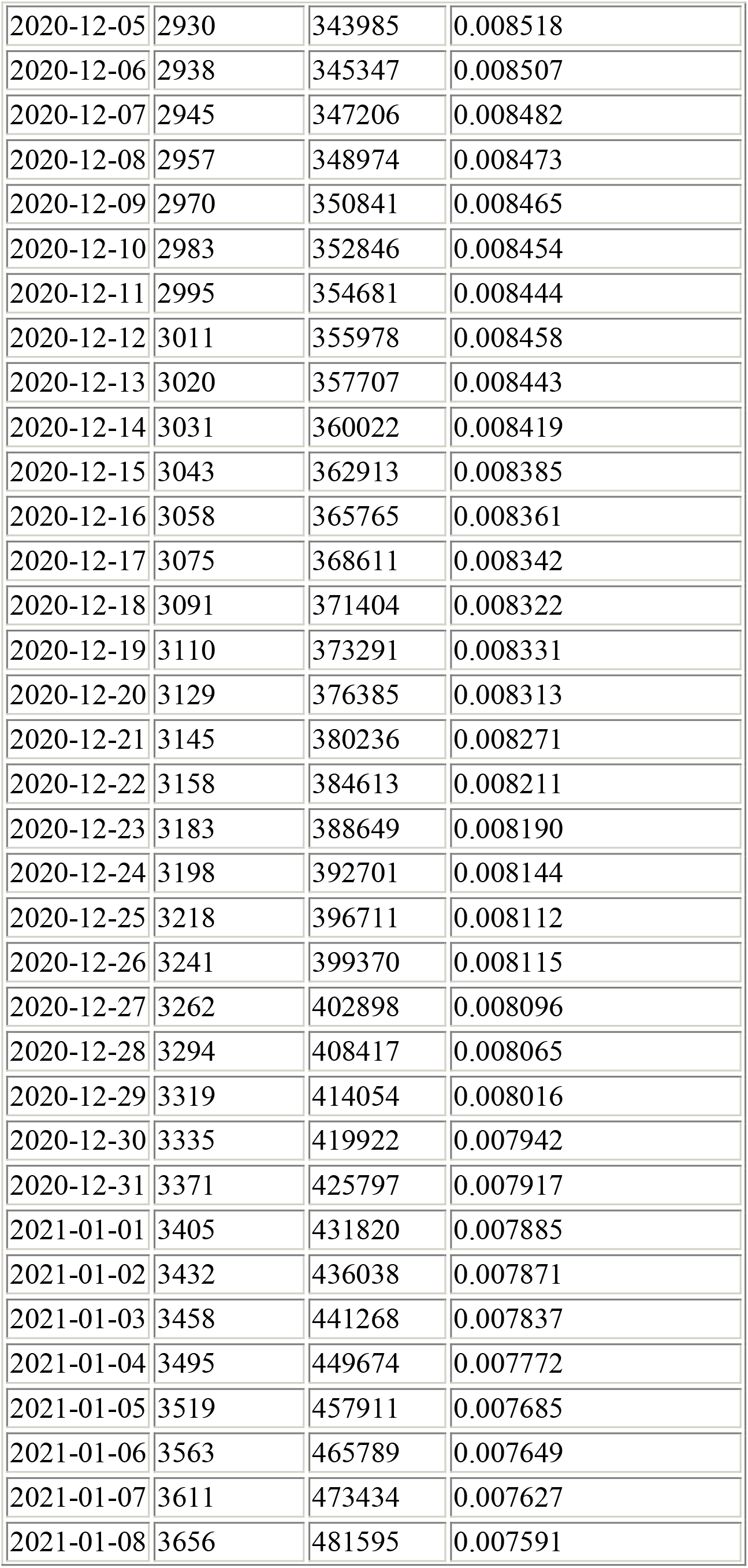
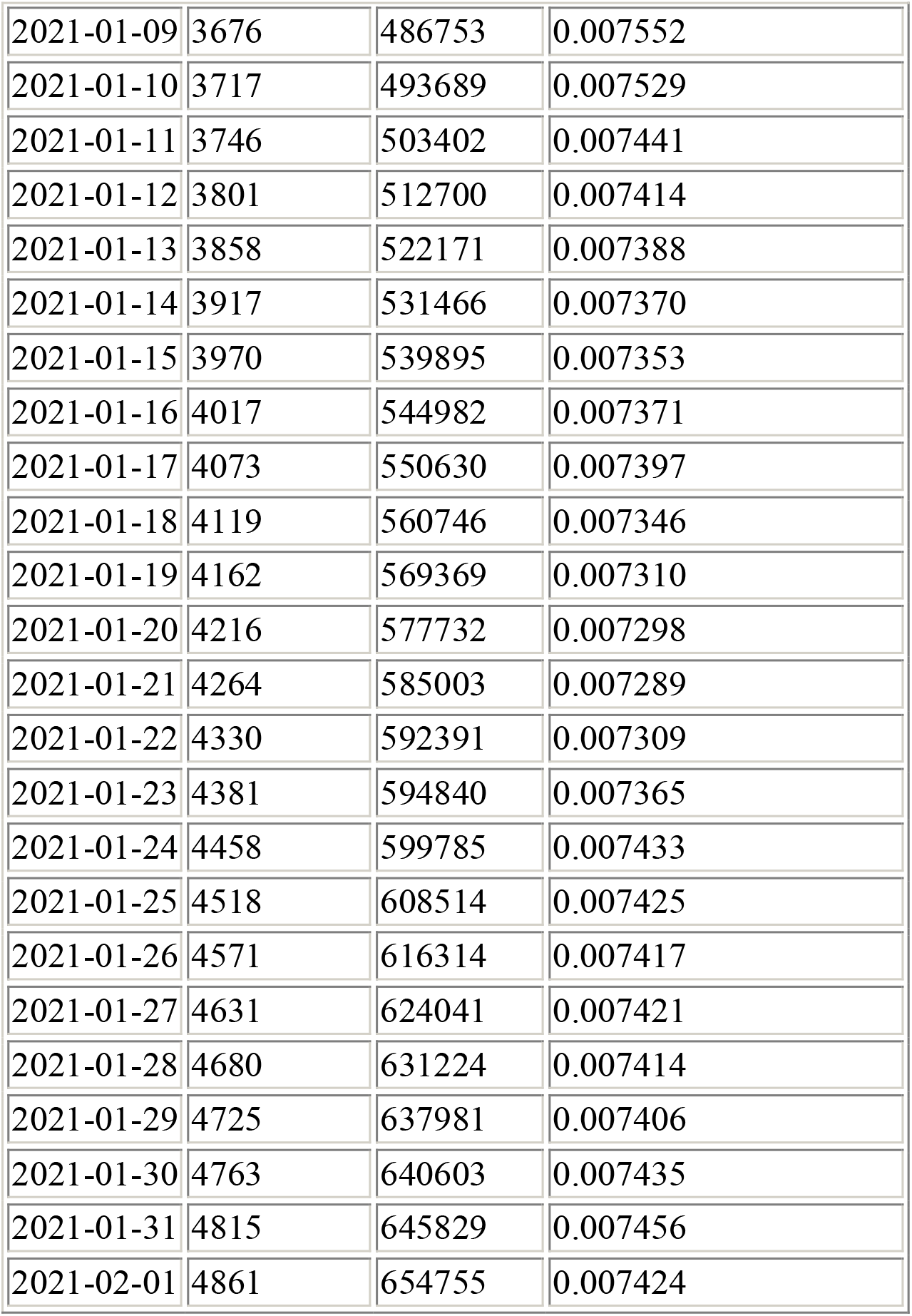

| Date | Total deaths | Total cases | Cumulative case fatality |
| --- | --- | --- | --- |
| 2020-11-01 | 2593 | 315051 | 0.008230 |
| 2020-11-02 | 2608 | 315840 | 0.008257 |
| 2020-11-03 | 2634 | 316685 | 0.008317 |
| 2020-11-04 | 2649 | 317454 | 0.008345 |
| 2020-11-05 | 2663 | 318024 | 0.008374 |
| 2020-11-06 | 2679 | 318715 | 0.008406 |
| 2020-11-07 | 2687 | 318927 | 0.008425 |
| 2020-11-08 | 2694 | 319461 | 0.008433 |
| 2020-11-09 | 2701 | 320180 | 0.008436 |
| 2020-11-10 | 2711 | 320855 | 0.008449 |
| 2020-11-11 | 2718 | 321620 | 0.008451 |
| 2020-11-12 | 2730 | 322456 | 0.008466 |
| 2020-11-13 | 2735 | 323217 | 0.008462 |
| 2020-11-14 | 2747 | 323508 | 0.008491 |
| 2020-11-15 | 2756 | 324134 | 0.008503 |
| 2020-11-16 | 2764 | 325007 | 0.008504 |
| 2020-11-17 | 2776 | 325836 | 0.008520 |
| 2020-11-18 | 2783 | 326631 | 0.008520 |
| 2020-11-19 | 2791 | 327402 | 0.008525 |
| 2020-11-20 | 2797 | 328176 | 0.008523 |
| 2020-11-21 | 2809 | 328598 | 0.008548 |
| 2020-11-22 | 2825 | 329360 | 0.008577 |
| 2020-11-23 | 2832 | 330319 | 0.008574 |
| 2020-11-24 | 2838 | 331176 | 0.008569 |
| 2020-11-25 | 2843 | 332255 | 0.008557 |
| 2020-11-26 | 2852 | 333342 | 0.008556 |
| 2020-11-27 | 2859 | 334373 | 0.008550 |
| 2020-11-28 | 2868 | 334955 | 0.008562 |
| 2020-11-29 | 2878 | 335979 | 0.008566 |
| 2020-11-30 | 2885 | 337232 | 0.008555 |
| 2020-12-01 | 2895 | 338431 | 0.008554 |
| 2020-12-02 | 2900 | 340019 | 0.008529 |
| 2020-12-03 | 2910 | 341471 | 0.008522 |
| 2020-12-04 | 2920 | 342985 | 0.008513 |
| 2020-12-05 | 2930 | 343985 | 0.008518 |
| 2020-12-06 | 2938 | 345347 | 0.008507 |
| 2020-12-07 | 2945 | 347206 | 0.008482 |
| 2020-12-08 | 2957 | 348974 | 0.008473 |
| 2020-12-09 | 2970 | 350841 | 0.008465 |
| 2020-12-10 | 2983 | 352846 | 0.008454 |
| 2020-12-11 | 2995 | 354681 | 0.008444 |
| 2020-12-12 | 3011 | 355978 | 0.008458 |
| 2020-12-13 | 3020 | 357707 | 0.008443 |
| 2020-12-14 | 3031 | 360022 | 0.008419 |
| 2020-12-15 | 3043 | 362913 | 0.008385 |
| 2020-12-16 | 3058 | 365765 | 0.008361 |
| 2020-12-17 | 3075 | 368611 | 0.008342 |
| 2020-12-18 | 3091 | 371404 | 0.008322 |
| 2020-12-19 | 3110 | 373291 | 0.008331 |
| 2020-12-20 | 3129 | 376385 | 0.008313 |
| 2020-12-21 | 3145 | 380236 | 0.008271 |
| 2020-12-22 | 3158 | 384613 | 0.008211 |
| 2020-12-23 | 3183 | 388649 | 0.008190 |
| 2020-12-24 | 3198 | 392701 | 0.008144 |
| 2020-12-25 | 3218 | 396711 | 0.008112 |
| 2020-12-26 | 3241 | 399370 | 0.008115 |
| 2020-12-27 | 3262 | 402898 | 0.008096 |
| 2020-12-28 | 3294 | 408417 | 0.008065 |
| 2020-12-29 | 3319 | 414054 | 0.008016 |
| 2020-12-30 | 3335 | 419922 | 0.007942 |
| 2020-12-31 | 3371 | 425797 | 0.007917 |
| 2021-01-01 | 3405 | 431820 | 0.007885 |
| 2021-01-02 | 3432 | 436038 | 0.007871 |
| 2021-01-03 | 3458 | 441268 | 0.007837 |
| 2021-01-04 | 3495 | 449674 | 0.007772 |
| 2021-01-05 | 3519 | 457911 | 0.007685 |
| 2021-01-06 | 3563 | 465789 | 0.007649 |
| 2021-01-07 | 3611 | 473434 | 0.007627 |
| 2021-01-08 | 3656 | 481595 | 0.007591 |
| 2021-01-09 | 3676 | 486753 | 0.007552 |
| 2021-01-10 | 3717 | 493689 | 0.007529 |
| 2021-01-11 | 3746 | 503402 | 0.007441 |
| 2021-01-12 | 3801 | 512700 | 0.007414 |
| 2021-01-13 | 3858 | 522171 | 0.007388 |
| 2021-01-14 | 3917 | 531466 | 0.007370 |
| 2021-01-15 | 3970 | 539895 | 0.007353 |
| 2021-01-16 | 4017 | 544982 | 0.007371 |
| 2021-01-17 | 4073 | 550630 | 0.007397 |
| 2021-01-18 | 4119 | 560746 | 0.007346 |
| 2021-01-19 | 4162 | 569369 | 0.007310 |
| 2021-01-20 | 4216 | 577732 | 0.007298 |
| 2021-01-21 | 4264 | 585003 | 0.007289 |
| 2021-01-22 | 4330 | 592391 | 0.007309 |
| 2021-01-23 | 4381 | 594840 | 0.007365 |
| 2021-01-24 | 4458 | 599785 | 0.007433 |
| 2021-01-25 | 4518 | 608514 | 0.007425 |
| 2021-01-26 | 4571 | 616314 | 0.007417 |
| 2021-01-27 | 4631 | 624041 | 0.007421 |
| 2021-01-28 | 4680 | 631224 | 0.007414 |
| 2021-01-29 | 4725 | 637981 | 0.007406 |
| 2021-01-30 | 4763 | 640603 | 0.007435 |
| 2021-01-31 | 4815 | 645829 | 0.007456 |
| 2021-02-01 | 4861 | 654755 | 0.007424 |

## Discussion

Although crude mortality rates were higher with the alpha variant strain, the case fatality rates were lower. Escalating crude mortality rates and decreasing case fatality rates can occur due to one or more of many mechanisms. Although there are many possible explanations for this, given the extensive free testing apparatus throughout Israel and lack of changes in the procedures of this apparatus, one obvious mechanism for this scenario would be to have a more infectious but not more deadly or even less deadly strain to emerge as the predominant one.

In theory a dramatic change in population vaccination level could have brought on changes in case fatality rate. However, the amount of vaccination in the population was not enough to significantly change the fatality of the disease in the critical period in which the alpha variant became predominant and statistical analyses were performed. The Israeli vaccination campaign began on December 20^th^ and by January 10^th^ less than 1% of both the general population and the population over 50 had been completely vaccinated with both shots in the two-shot regimen (11). While there is some protective effect of a single vaccination, the cumulative incidence curves for patients with a single vaccination only begin to diverge after two weeks (12). Thus, dates before mid-January were chosen for statistical analysis to avoid the potential confounding effect of single-dose vaccination.

A possible explanation of our findings is that alpha variant infected more people over time in Israel and the UK, but once infected, these people were no more likely to die than people infected with the wild-type virus. These findings would contradict the hypothesis that infection with alpha variant is associated with an increased risk of death compared to infection with non-alpha variant strains (13) if the people infected by the alpha variant had the same underlying risk of death. There is the theoretical possibility that the period in which the alpha variant rose saw a shift in the demographics of cases, whether in terms of age or co-morbidities, which changed the case fatality. However, surveillance data suggests that the b117 rose rapidly in all age populations until mid-January (4).

Our evidence is based on the data from Israel, but publicly available data reveals that in many countries, including the UK, fatality rates are not rising above levels in December 2020, although alpha variant has widely spread (14). UK case fatality rates have been higher than Israeli ones as Israel has a younger population than the UK. Israel is the world leader in COVID-19 vaccinations, but they significantly affected infections and mortality in March 2021.

In conclusion, although alpha variant appears more transmissible, our findings suggest that it is not more deadly once a patient is infected. To combat the COVID pandemic as the alpha variant becomes the dominant strain, preventing deaths should focus on preventing transmission.

## Data Availability

The data for this article was made from the original publically available data of the Israel Ministry of Health online, and is cited. In some cases, data has been processed from this original data.

## Data availability

Data and code are available upon request.

## Acknowledgements

The MaxCor lab is the recipient of funds from the Israel Institute for Health Policy Research with grant number 2020/442. The funder had no direct role in the development of the report or analysis and no involvement in the publication process.

## References

1. Eurosurveillance Editorial Team, 2021. Updated rapid risk assessment from ECDC on the risk related to the spread of new SARS-CoV-2 variants of concern in the EU/EEA–first update. Eurosurveillance, 26(3), 2021 p. 2101211.

2. Davies, N.G., Jarvis, C.I., Edmunds, W.J., Jewell, N.P., Diaz-Ordaz, K. and Keogh, R.H., 2021. Increased mortality in community-tested cases of SARS-CoV-2 lineage B. 1.1. 7. Nature, 593(7858), pp. 270–274.

3. Galloway SE, Paul P, MacCannell DR, et al. Emergence of SARS-COV-2 B.1.1.7 Lineage-United States, December 29^th^, 2020-January 12th, 2021. MMWR Morb. Mortal Wkly Rep 2021; 70:95–99.

4. Munitz, A., Yechezkel, M., Dickstein, Y., Yamin, D. and Gerlic, M., 2021. The rise of SARS-CoV-2 variant B. 1.1. 7 in Israel intensifies the role of surveillance and vaccination in elderly. Cell Reports Medicine, 2021.

5. Volz, E., Mishra, S., Chand, M., Barrett, J.C., Johnson, R., Geidelberg, L., Hinsley, W.R., Laydon, D.J., Dabrera, G., O’Toole, Á. and Amato, R., 2021. Assessing transmissibility of SARS-CoV-2 lineage B. 1.1. 7 in England. Nature, 2021, pp. 1–17.

6. Iacobucci, G., Covid-19: New UK variant may be linked to increased death rate, early data indicate. BMJ, 2021, 372(230), p. n230.

7. Challen, R., Brooks-Pollock, E., Read, J.M., Dyson, L., Tsaneva-Atanasova, K. and Danon, L., Risk of mortality in patients infected with SARS-CoV-2 variant of concern 202012/1: matched cohort study. BMJ, 2021, 372.

8. Grint, D.J., Wing, K., Williamson, E., McDonald, H.I., Bhaskaran, K., Evans, D., Evans, S.J., Walker, A.J., Hickman, G., Nightingale, E. and Schultze, A., Case fatality risk of the SARS-CoV-2 variant of concern B.1.1.7 in England, November 16th to February 5th. Eurosurveillance, 2021, 26(11), p. 2100256.

9. Frampton, D., Rampling, T., Cross, A., Bailey, H., Heaney, J., Byott, M., Scott, R., Sconza, R., Price, J., Margaritis, M. and Bergstrom, M., Genomic characteristics and clinical effect of the emergent SARS-CoV-2 B. 1.1. 7 lineage in London, UK: a whole-genome sequencing and hospital-based cohort study. The Lancet Infectious Diseases. 2021. Published online April 2021. Available from https://www.thelancet.com/pdfs/journals/laninf/PIIS1473-3099(21)00170-5.pdf

10. Temsah, Mohamad-Hani, et al. SARS-CoV-2 B.1.1.7 UK Variant of Concern Lineage-Related Perceptions, COVID-19 Vaccine Acceptance and Travel Worry Among Healthcare Workers. May 26th, 2021, Front. Public Health, vol. 9, p. doi: 10.3389/fpubh.2021.686958.

11. Israel Ministry of Heath, “המתחסנים גילאי” file published online in information databases presented the Israeli Ministry of Communication’s at (https://data.gov.il/dataset/covid-19), specific dataset last accessed at (https://data.gov.il/dataset/covid-19/resource/57410611-936c-49a6-ac3c-838171055b1f) on 2/8/2021

12. Dagan, N., Barda, N., Kepten, E., Miron, O., Perchik, S., Katz, M.A., Hernán, M.A., Lipsitch, M., Reis, B. and Balicer, R.D. BNT162b2 mRNA Covid-19 vaccine in a nationwide mass vaccination setting. New England Journal of Medicine, 384(15), pp. 1412-1432. DOI: 10.1056/NEJMoa2101765

13. Peter Horby, Catherine Huntley, Nick Davies, John Edmunds, Niel Ferguson, Granam Medley, Calum Semple, NERVTAG paper on COVID-19 Variant of Concern B.1.1.7 Available from: https://www.gov.uk/government/publications/nervtag-paper-on-covid-19-variant-of-concern-b117; Accessed May 5th, 2021.

14. Hannah Ritchie, Esteban Ortiz-Ospina, Diana Beltekian, Edouard Mathieu, Joe Hasell, Bobbie Macdonald, Charlie Giattino, Cameron Appel, Lucas Rodés-Guirao and Max Roser, Mortality Risk of COVID-19. Available from https://ourworldindata.org/mortality-risk-covid?country=ISR∼GBR; Accessed May 5th, 2021.

